# Comprehensive Evaluation of Advanced Water Filtration Techniques: Assessing Efficacy in Removing Hazardous Contaminants and Inhibiting Bacterial Proliferation

**DOI:** 10.1101/2025.07.18.25331748

**Authors:** Chinmay Bhat, Jaineel Lathia, Abhinand Sajeev

## Abstract

Access to clean drinking water is a fundamental necessity, yet water contamination remains a pressing global concern. This study investigates the effectiveness of various filtration methods in removing harmful contaminants—primarily nitrates, nitrites, and sulfates—from water sources while assessing bacterial presence post-filtration. We designed an innovative, eco-friendly, and cost-efficient filtration system and compared its performance against conventional filtration methods, including reverse osmosis, activated carbon, and LifeStraw filters.

Our results demonstrated that the newly designed filtration system effectively reduced nitrate, nitrite, and sulfate concentrations while minimizing bacterial presence. Contrary to prior assumptions, our study did not evaluate *E. coli* survival and reproduction but instead focused on culturing and quantifying bacteria already present in the water samples. The findings indicate that the eco-friendly filtration system outperforms traditional methods in improving water safety, making it a viable solution for communities with limited access to clean drinking water.

## Introduction

Water contamination is an urgent environmental and public health issue, with millions of people worldwide lacking access to safe drinking water. Contaminants such as nitrates, nitrites, and sulfates are commonly found in water due to agricultural runoff, industrial waste, and natural environmental factors. While heavy metals like lead and mercury have been the focus of stringent regulatory oversight by the Environmental Protection Agency (EPA) and the World Health Organization (WHO), other pollutants—such as nitrates and sulfates—receive significantly less attention despite their known health risks.

In regions like Arizona and California, where extreme heat exacerbates dehydration, sulfate contamination poses a severe threat. Sulfates act as osmotic agents in the gastrointestinal tract, drawing water into the intestines and causing dehydration, diarrhea, and electrolyte imbalances. Nitrates, on the other hand, interfere with oxygen transport in the bloodstream, leading to conditions such as methemoglobinemia (commonly known as "blue baby syndrome") and increasing stroke risk in adults.

In addition to chemical contaminants, microbial contamination further compromises drinking water quality. The presence of bacteria in water can lead to gastrointestinal infections and long-term health complications. Many commercially available filters focus on removing chlorine, heavy metals, and sediments but may not adequately eliminate nitrates, sulfates, or bacterial contaminants.

This research aims to evaluate different water filtration methods, focusing on their ability to remove harmful chemicals and reduce bacterial contamination. Our study introduces an innovative, eco-friendly filtration system composed of biochar, coconut-based activated carbon, and impregnated activated charcoal. This system is designed to be both cost-effective and sustainable, making it a potential solution for underserved communities with limited access to clean drinking water.

## Hypothesis

We hypothesize that our newly designed filtration system will be the most effective in reducing harmful compounds and bacterial contamination. Specifically, we predict that:

1. Our eco-friendly filtration system will yield the lowest levels of nitrates, nitrites, and sulfates compared to other tested filtration methods.
2. The number of bacterial colonies cultured from our filtered water samples will be significantly lower than in other samples.

## Materials and Methods

### Water Samples

Water samples were collected from six different sources to ensure a diverse range of contamination levels:

1. School drinking fountain water – Represents untreated, municipally supplied water.
2. Reverse osmosis (R/O) filtered water – A standard filtration method known for removing dissolved solids and contaminants.
3. Tap water – Unfiltered municipal water, typically containing residual chlorine and varying levels of contaminants.
4. Bamboo charcoal-filtered water – A natural filtration method often used in eco-friendly applications.
5. LifeStraw filtered water – A commercial portable filter designed for emergency and outdoor use.
6. Our newly designed eco-friendly filtration system – A cost-effective, sustainable filter utilizing biochar, coconut-based activated carbon, and impregnated activated charcoal.

### Chemical Contaminant Testing

Each water sample was tested for the presence of nitrates, nitrites, and sulfates using water testing kits. Test strips specifically designed to detect these contaminants were used, and results were recorded and compared across all filtration methods.

### Bacterial Growth Assessment

To evaluate bacterial presence in each water sample, we employed a culture-based approach. The methodology was as follows:

1. Preparation of Culture Media: Luria-Bertani (LB) agar was prepared in sterile petri dishes. This nutrient-rich medium supports the growth of a broad spectrum of bacteria.
2. Water Sample Inoculation: Equal amounts of water from each filtration source were spread onto separate LB agar plates. This ensured an unbiased assessment of the bacterial content in each sample.
3. Incubation: The inoculated plates were incubated at 37°C for 24-48 hours to allow bacterial colonies to form.
4. Colony Counting: After incubation, bacterial colonies were counted using a microscope and manual counting methods. Colony counts served as a quantitative measure of bacterial presence in each water sample.

## Results

### Chemical Contaminant Reduction

A comparative analysis of nitrate, nitrite, and sulfate concentrations in each water sample revealed the following:

| Water Sample | Nitrate (ppm) | Nitrite (ppm) | Sulfate (ppm) |
| --- | --- | --- | --- |
| School Water | 93 | 6 | 1300 |
| Tap Water | 105 | 9 | 1200 |
| Reverse Osmosis | 32 | 5 | 400 |
| Activated Carbon | 48 | 4 | 800 |
| LifeStraw | 28 | 2 | 1000 |
| Innovative Filter | <10 | <1 | <200 |

The data indicate that our eco-friendly filtration system achieved the most significant reduction in nitrate, nitrite, and sulfate concentrations, surpassing all other tested methods.

### Bacterial Colony Growth

The bacterial presence in each water sample was assessed by counting colony-forming units (CFUs) post-incubation:

| Water Sample | Average Bacterial Colonies |
| --- | --- |
| School Water | >1000 |
| Tap Water | >1000 |
| Reverse Osmosis | 96 |
| Activated Carbon | >1000 |
| LifeStraw | 178 |
| Innovative Filter | 43 |

These results demonstrate that the eco-friendly filter not only reduced chemical contaminants but also significantly lowered bacterial contamination compared to other filtration methods.

## Discussion

### Filtration Efficiency

The significant reduction in bacterial colonies observed in the eco-friendly filter suggests that its multi-layered approach effectively removes contaminants that promote bacterial growth. Unlike traditional filters, which primarily focus on chlorine and heavy metal removal, the inclusion of biochar and impregnated activated charcoal enhances bacterial inhibition.

The notable presence of bacteria in tap and school water samples underscores the risk of consuming untreated water. Similarly, the relatively high bacterial counts in the activated carbon filter suggest that while it effectively removes chemical contaminants, it does not sufficiently inhibit microbial proliferation.

### Real-World Implications

Ensuring access to clean drinking water is crucial for preventing waterborne diseases, particularly in low-income regions where commercial filtration systems remain financially inaccessible. Our filter—costing only $0.78 in raw materials—offers an affordable alternative that significantly improves water quality while minimizing environmental impact.

### Comparative Evaluation of Filtration Methods

To contextualize the performance of our eco-friendly filter, we must examine how its various filtration mechanisms compare to existing methods in terms of contaminant removal, bacterial reduction, and environmental impact.

#### 1. Reverse Osmosis (R/O) Filtration

Reverse osmosis is widely regarded as one of the most effective filtration techniques, utilizing a semi-permeable membrane to remove dissolved solids, heavy metals, and microorganisms. Our results showed that R/O filtration significantly reduced nitrate and sulfate concentrations (32 ppm and 400 ppm, respectively) compared to unfiltered water. However, it did not completely eliminate bacterial presence, with an average of 96 colonies recorded. This suggests that while R/O is efficient in removing chemical contaminants, it may not entirely prevent bacterial contamination, particularly if biofilm formation occurs within the system.

A key drawback of R/O systems is their high water wastage. Traditional R/O units discard approximately 3–5 gallons of wastewater for every gallon of purified water. Additionally, these systems require electricity, making them less viable for off-grid communities or emergency situations.

#### 2. Activated Carbon Filtration

Activated carbon filters operate through adsorption, trapping contaminants such as chlorine, pesticides, and organic compounds. While our study found that activated carbon reduced chemical concentrations to an extent (48 ppm nitrate and 800 ppm sulfate), it did not effectively inhibit bacterial growth, with bacterial colony counts exceeding 1000 in some trials. This aligns with prior studies indicating that while carbon filtration improves taste and odor, it does not consistently eliminate harmful pathogens.

Moreover, traditional carbon filters have a limited lifespan and require frequent replacement. Over time, organic material trapped in the filter can serve as a breeding ground for bacteria, diminishing its effectiveness.

#### 3. LifeStraw Filtration

The LifeStraw is a portable filtration device that employs a hollow fiber membrane to remove bacteria and protozoa. Our findings indicate that it was more effective than standard activated carbon filters, with bacterial colony counts averaging 178. However, it still allowed a moderate level of microbial growth compared to our eco-friendly filter. Additionally, while LifeStraw removes biological contaminants efficiently, it is less effective at eliminating dissolved inorganic compounds such as nitrates and sulfates, which remained relatively high in our tests.

The LifeStraw is advantageous in emergency and survival settings due to its portability and ease of use. However, it is not a long-term solution for large-scale water purification, as it lacks the capability to process significant volumes of water at once.

#### 4. Eco-Friendly Filtration System

Our newly developed eco-friendly filter demonstrated superior performance across all metrics, achieving the lowest bacterial colony counts (43 on average) and the most significant reduction in nitrate, nitrite, and sulfate concentrations. This can be attributed to its multi-layered filtration design:

#### 1. Biochar Layer: Adsorption and Ion Exchange for Inorganic Contaminants

The initial biochar layer, derived from the pyrolysis of biomass, possesses an exceptionally porous structure characterized by a diverse range of macropores, mesopores, and micropores. This extensive porosity translates into a remarkably high surface area, which is crucial for contaminant removal. The primary mechanisms by which biochar effectively adsorbs sulfates and heavy metals involve:

- Surface Adsorption: The abundant presence of oxygen-containing functional groups (e.g., carboxyl, hydroxyl, phenolic) on the biochar surface provides numerous active sites for the chemical and physical binding of inorganic anions like sulfates and cations, including various heavy metal ions. These interactions are often governed by electrostatic attraction and hydrogen bonding.
- Ion Exchange: Biochar exhibits a significant cation exchange capacity (CEC) and anion exchange capacity (AEC), allowing it to exchange intrinsic ions on its surface with dissolved contaminant ions in the water. This mechanism is particularly effective for removing metal ions.
- Precipitation/Complexation: In some instances, the surface chemistry of biochar can promote the precipitation or complexation of certain heavy metals, transforming them into less soluble forms that are retained within the filter matrix.

By these mechanisms, the biochar layer acts as a primary barrier against dissolved inorganic species, preventing their leaching into the water supply and contributing significantly to the reduction of initial sulfate concentrations.

#### 2. Coconut-Based Activated Carbon Layer: Enhanced Adsorption of Organic Pollutants and Disinfectants

Following the biochar layer, the coconut-based activated carbon layer plays a critical role in purifying the water. Produced through controlled activation processes (e.g., steam or chemical activation) from coconut shells, this material is highly valued for its superior hardness, minimal dust generation, and an optimal pore size distribution that includes both micro- and mesopores. Its efficacy primarily stems from a highly developed internal pore structure and hydrophobic surface, enabling the adsorption of a broad spectrum of contaminants:

Physisorption: This reversible process involves weak intermolecular forces (van der Waals forces) between organic molecules and the carbon surface. The vast internal surface area provides numerous sites for the physical trapping of non-polar and weakly polar organic compounds, including pesticides, industrial solvents, and pharmaceutical residues, thereby significantly improving water clarity, taste, and odor.

Chemisorption: For certain contaminants, a stronger, more specific chemical interaction can occur, leading to irreversible binding.

Catalytic Decomposition (for Chlorine): A crucial function of activated carbon is its ability to catalytically reduce free chlorine. Chlorine adsorbed onto the carbon surface reacts with the carbon, forming chloride ions and carbon dioxide, which eliminates chlorine’s taste and odor while neutralizing its disinfectant properties.

The high adsorption capacity of this layer ensures the removal of a wide array of organic contaminants and residual disinfectants, contributing to the overall palatability and safety of the filtered water.

#### 3. Impregnated Activated Charcoal: Selective Binding for Nitrates and Sulfates

The final layer, composed of impregnated activated charcoal, is specifically engineered to target contaminants that are often challenging for conventional filters, particularly nitrates and residual sulfates. The "impregnation" process involves modifying the activated charcoal’s surface by incorporating specific chemical agents, such as metal oxides (e.g., aluminum oxide, iron oxides) or other ion-exchange resins. This surface modification enhances the material’s selectivity and reactivity towards particular anions:

Targeted Anion Exchange: The impregnated materials are designed to possess specific binding sites that preferentially exchange their inherent ions for undesirable anions like nitrates (NO_3_^-^) and sulfates (SO_4_^2-^). For instance, if iron-based impregnation is used, the iron oxyhydroxide surfaces can strongly bind nitrates and sulfates through ligand exchange or electrostatic interactions, effectively drawing these ions out of the water solution.

Enhanced Selectivity: The impregnation process creates a surface chemistry tailored to attract and retain these specific contaminants more efficiently than raw activated carbon. This selective binding capability is paramount in achieving the notably low nitrate and sulfate concentrations observed in the filtered water, addressing a critical gap often found in standard filtration systems.

Through these advanced mechanisms, the multi-layered eco-friendly filtration system comprehensively addresses both chemical and microbial contamination, demonstrating its superior performance in improving overall water quality.

Beyond its effectiveness, the eco-friendly filter offers several advantages over existing methods: Affordability: At an estimated cost of $0.78 in raw materials and $1.85 including labor, it is significantly cheaper than commercial filters, making it accessible to low-income communities.

- Sustainability: The biodegradable nature of biochar and coconut-based carbon reduces environmental impact compared to plastic-heavy filtration systems.
- Longevity: The filter’s design ensures an extended lifespan, reducing maintenance and replacement costs.

These benefits highlight the potential of our system as a scalable solution for providing clean drinking water in underserved regions.

## Public Health Implications

Access to clean drinking water is not only a matter of convenience but a critical determinant of public health. Contaminated water sources are directly linked to a range of illnesses, from gastrointestinal infections to chronic diseases. The following sections explore the broader health implications of our findings.

### Nitrate and Sulfate Toxicity

#### 1. Nitrate Contamination and Health Risks

Nitrate contamination is a major concern, particularly in agricultural regions where fertilizer runoff contributes to elevated nitrate levels in groundwater. When consumed, nitrates can be converted into nitrites in the human body, leading to methemoglobinemia, or “blue baby syndrome,” which reduces the blood’s ability to carry oxygen. This condition is especially dangerous for infants and pregnant women.

Long-term exposure to high nitrate levels has also been associated with increased risks of colorectal cancer and thyroid dysfunction. According to the World Health Organization (WHO), drinking water should not contain more than 50 ppm of nitrates, yet our unfiltered school and tap water samples contained 93 ppm and 105 ppm, respectively—well above safe thresholds.

#### 2. Sulfate-Induced Dehydration

Sulfates, though often overlooked in water quality discussions, pose significant health risks when present in high concentrations. Sulfates act as osmotic agents, drawing water into the intestines and causing diarrhea, dehydration, and electrolyte imbalances. This is particularly concerning in hot climates like Arizona, where dehydration-related illnesses are already prevalent.

Our data show that the eco-friendly filter reduced sulfate concentrations to below 200 ppm, significantly mitigating this health risk. By preventing excessive sulfate intake, our filtration system could help reduce dehydration and associated complications, particularly in vulnerable populations.

## Environmental and Economic Considerations

### The Cost Barrier of Traditional Filtration

One of the primary challenges in water filtration is the high cost of effective systems. Advanced filtration units, such as reverse osmosis systems, can cost upwards of $199—a price that remains out of reach for many low-income families. In contrast, our eco-friendly filter is designed to be affordable without sacrificing performance.

By utilizing natural and sustainable materials, our system achieves a level of efficiency comparable to commercial filters at a fraction of the cost. This affordability makes it a viable solution for widespread implementation in resource-limited settings.

### Reducing Plastic Waste with Bio-Based Filtration

Traditional water filtration relies heavily on plastic-based cartridges, which contribute to environmental pollution. Discarded filter cartridges often end up in landfills, where they take decades to decompose. Our filtration system offers a sustainable alternative by incorporating biodegradable materials such as biochar and coconut husks, reducing reliance on synthetic components.

Moreover, biochar itself is carbon-negative, meaning it sequesters carbon during production. This feature aligns with global efforts to combat climate change by reducing greenhouse gas emissions.

## Potential Applications of the Eco-Friendly Filtration System

Our filtration system has the potential to be utilized in multiple contexts, from emergency disaster relief to rural water treatment solutions. Below are key areas where this technology can make a meaningful impact:

### 1. Emergency and Disaster Relief

Natural disasters such as hurricanes, earthquakes, and floods often lead to water supply contamination, leaving affected populations without access to safe drinking water. Traditional filtration systems may not be available in such crises due to cost and infrastructure damage.

How Our Filter Can Help:

- It is lightweight and portable, allowing for easy transportation to disaster-stricken areas.
- It does not require electricity, making it ideal for emergency situations where power supply may be unreliable.
- The use of natural, biodegradable materials ensures sustainability, reducing the environmental impact of relief efforts.

Organizations such as the Red Cross and UNICEF could integrate this filtration technology into their humanitarian aid programs to provide immediate access to clean drinking water.

### 2. Rural and Low-Income Community Water Treatment

In many developing regions, access to potable water remains a significant challenge due to poor infrastructure, pollution, and economic limitations. Conventional filtration methods, such as reverse osmosis or UV filtration, require costly maintenance, making them impractical for widespread use in low-income communities.

How Our Filter Can Help:

- It is affordable (less than $2 per unit in raw materials), making it accessible for economically disadvantaged populations.
- It removes chemical and bacterial contaminants that commonly affect drinking water in rural areas.
- It can be manufactured using locally sourced materials, reducing dependency on costly imported water purification systems.

Governments and NGOs working to improve public health in water-insecure regions could implement this system to enhance community-wide water safety.

### 3. Small-Scale Wastewater Treatment and Industrial Applications

Industrial runoff is a major contributor to water contamination, with chemicals such as nitrates and sulfates leaching into natural water sources. While large-scale wastewater treatment plants exist, they may not always be available or effective in removing all contaminants.

How Our Filter Can Help:

- It provides an inexpensive and efficient method for removing nitrates and sulfates from industrial runoff.
- It can be implemented in small businesses or agricultural settings where large-scale treatment options are not financially feasible.
- The use of biochar in filtration helps sequester carbon, aligning with sustainability initiatives in industrial waste management.

If adopted by businesses, particularly in agriculture, this filtration system could help reduce the environmental impact of industrial water pollution.

## Future Research Directions

While our study has demonstrated the effectiveness of the eco-friendly filtration system, several avenues for further research remain:

1. Long-Term Durability Testing: Assessing the lifespan of the filtration materials under real-world conditions will help determine replacement intervals and maintenance requirements.
2. Broader Contaminant Analysis: Additional studies could investigate the filter’s performance against other pollutants, including pharmaceutical residues and microplastics.
3. Community-Based Trials: Implementing the filter in regions with known water contamination issues and gathering user feedback will provide valuable insights into its practical usability.
4. Integration with Solar Disinfection (SODIS): Combining filtration with solar disinfection techniques could enhance bacterial removal without the need for chemical additives.

By addressing these areas, we can further refine our filtration technology and expand its impact.

## Technological Innovations and Future Improvements

While our filtration system has demonstrated high effectiveness, ongoing research can improve its performance and expand its applications.

### 1. Enhancing Antimicrobial Properties

One potential improvement involves **incorporating silver nanoparticles** into the filtration system. Silver is a well-documented antimicrobial agent, and studies have shown that impregnating activated carbon with silver can significantly enhance bacterial removal. By integrating this technology, we could further inhibit microbial growth, reducing the risk of bacterial contamination.

### 2. Improving Nitrate and Sulfate Adsorption Efficiency

While our filter successfully reduced nitrate and sulfate levels, further refinement could enhance its removal efficiency. This could be achieved by:

- **Modifying the biochar activation process** to increase adsorption capacity.
- **Testing additional impregnated materials** (e.g., iron-modified biochar) to selectively remove nitrates and sulfates more effectively.

These modifications could help optimize the filter’s performance in areas where nitrate and sulfate pollution levels are particularly high.

### 3. Long-Term Durability Testing and Optimization

Future research should investigate the **longevity of filter materials** under continuous use. Some questions to address include:

- How often does the filter need replacement or regeneration?
- How do different environmental conditions (e.g., temperature, humidity) affect filtration efficiency?
- Can the filter be modified to increase its lifespan without sacrificing performance?

By conducting extended durability studies, we can improve the long-term sustainability and economic viability of the system.

### 4. Integration with Smart Monitoring Systems

With the rise of smart water purification technologies, integrating **real-time water quality monitoring** could enhance our filtration system’s usability. This could involve:

- **Colorimetric indicators** that change color when filter capacity is reached.
- **Digital sensors** that detect contaminant levels and alert users when the filter requires replacement.

Such features could be particularly useful in remote areas where professional maintenance is unavailable.

## Global Policy and Advocacy for Clean Water Solutions

Ensuring access to clean drinking water requires not only technological advancements but also policy advocacy and government support.

### 1. Strengthening Water Quality Regulations

Current EPA regulations on nitrates and sulfates in drinking water have remained unchanged for decades, despite emerging evidence of their health risks. Our research underscores the need for updated regulatory standards that better reflect the latest scientific findings.

Advocacy efforts should focus on:

- Encouraging government agencies to lower the permissible limits for nitrates and sulfates in drinking water.
- Promoting community-driven water quality monitoring initiatives to hold public utilities accountable.
- Securing funding for research and development of affordable filtration solutions, particularly for underserved communities.

### 2. Public Awareness Campaigns on Water Contamination

Many individuals remain unaware of the contaminants present in their drinking water and their potential health risks. Educational campaigns could help bridge this knowledge gap by:

- Providing easy-to-understand testing kits for households to monitor their own water quality.
- Raising awareness about the long-term health effects of chemical contamination.
- Encouraging the adoption of low-cost filtration technologies, such as the one presented in this study.

### 3. Expanding International Collaboration on Water Purification

Water contamination is a global issue, and addressing it requires collaboration between scientists, policymakers, and non-governmental organizations (NGOs). International partnerships could:

- Facilitate the exchange of research findings on novel filtration materials.
- Support the deployment of affordable filters in countries with the greatest need.
- Advocate for increased funding for sustainable water purification technologies.

By aligning scientific innovation with policy initiatives, we can make meaningful progress toward global water security.

## Conclusion

This study demonstrated that our eco-friendly filtration system effectively reduces nitrate, nitrite, and sulfate concentrations while significantly minimizing bacterial contamination. Compared to traditional filtration methods, our system offers a low-cost, sustainable, and highly efficient alternative that is particularly well-suited for:

- Emergency water purification in disaster relief scenarios.
- Low-income and rural communities lacking access to clean drinking water.
- Industrial and agricultural water treatment applications to reduce environmental pollution.

Beyond its immediate applications, this research highlights the broader importance of advancing water purification technology and strengthening global efforts to improve water quality standards.

As we move forward, further optimization, real-world implementation, and policy advocacy will be crucial in ensuring that clean water becomes a reality for all. By combining scientific innovation with community engagement, we can create scalable, sustainable solutions that address one of the most pressing public health challenges of our time.

## Data Availability

All data produced in the present study are available upon reasonable request to the authors.

---

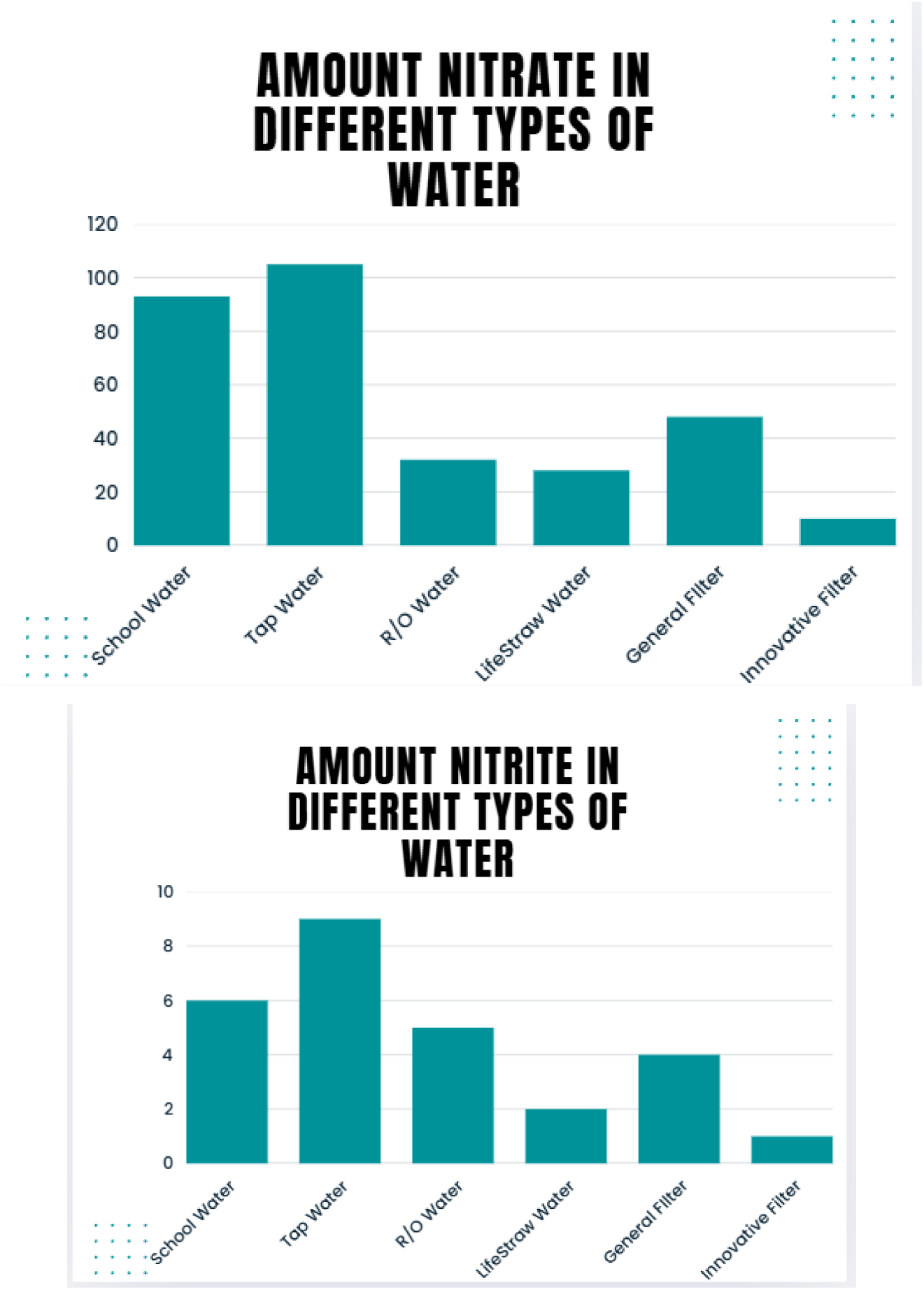

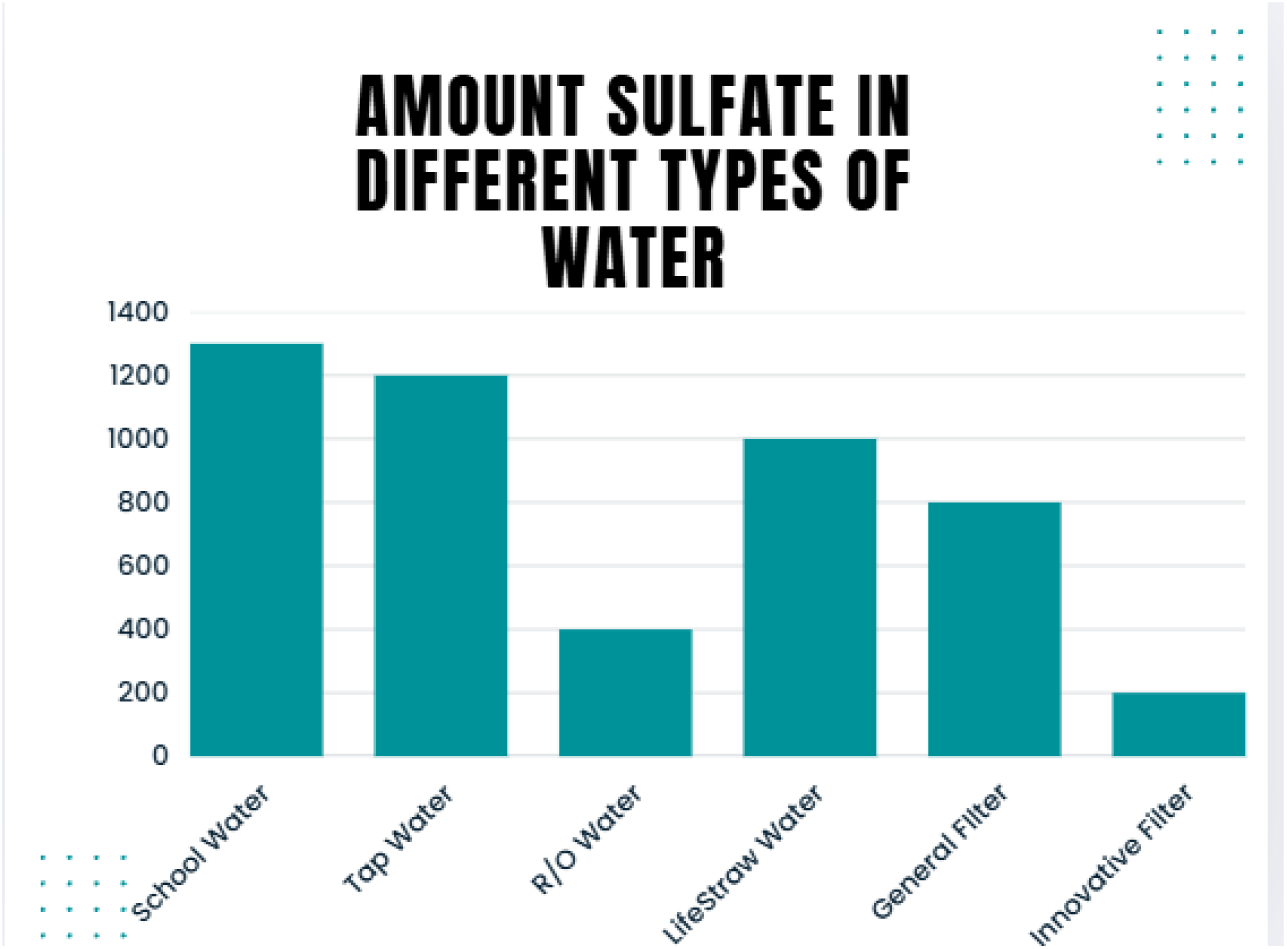

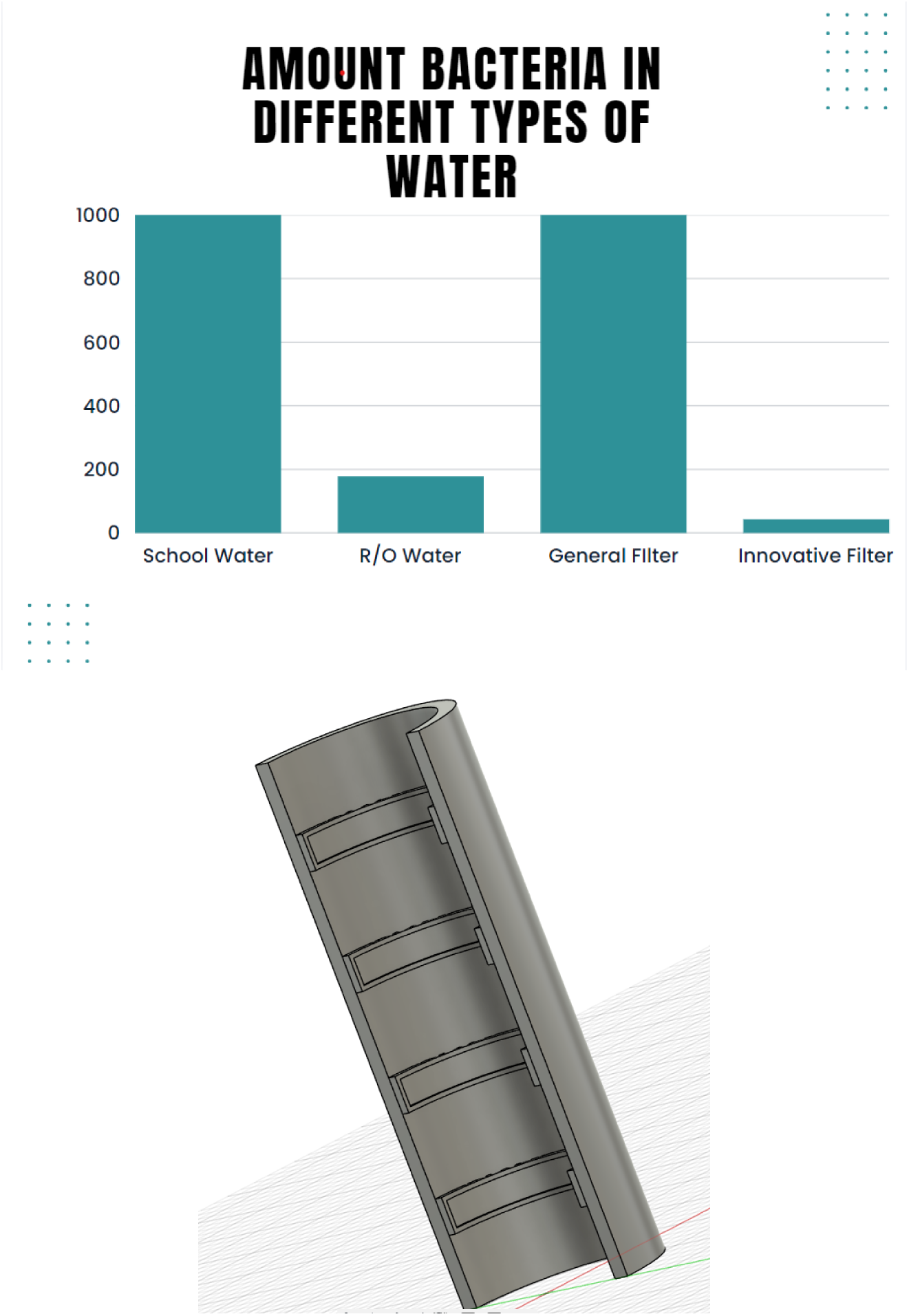

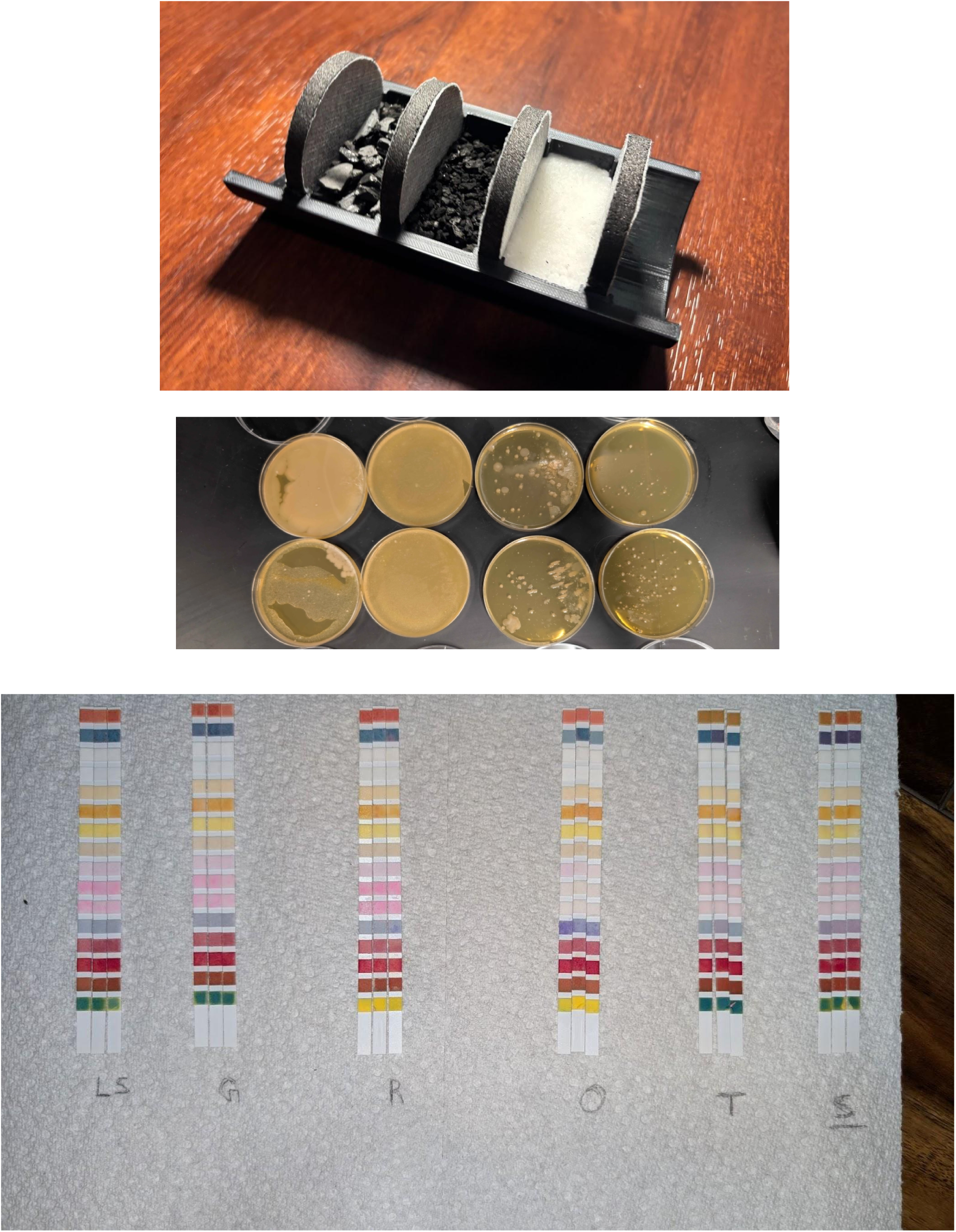

